# Procedural Pain, Rather Than Donor-Perceived Needle Quality, Is Associated With Willingness to Donate Again: A Multicentre Cross-Sectional Study in Bangladesh

**DOI:** 10.64898/2026.07.13.26357726

**Authors:** Ashraful Hoque, Maruful Rahman, Sushanta Kumar Basak, ABM Al Mamun

**Affiliations:** Department of Transfusion Medicine, Comilla Medical College Hospital, Cumilla, Bangladesh; Health System Performance Management, Ministry of Health, Government of Saskatchewan, Canada; Department of Transfusion Medicine, Faridpur Medical College Hospital, Faridpur, Bangladesh; Department of Transfusion Medicine, Dhaka Medical College Hospital, Dhaka, Bangladesh

**Keywords:** blood donation, donor retention, procedural pain, venepuncture, donor satisfaction, Bangladesh, transfusion medicine

## Abstract

**Background:** Retaining repeat blood donors is essential for maintaining a safe and sustainable blood supply. While pain during venepuncture has been linked to lower donor return, the influence of donor-perceived needle quality on future donation remains unclear, particularly in low- and middle-income countries. This study examined whether perceived needle quality was associated with willingness to donate again among blood donors in Bangladesh.

**Methods:** We conducted a cross-sectional analytical study among 100 consecutively recruited whole-blood donors from participating blood donation centres in Bangladesh. Participants completed a structured questionnaire assessing demographic characteristics, donation history, procedural pain using a 10-point Visual Analogue Scale (VAS), pre-donation fear, perceived needle quality, vasovagal symptoms, overall satisfaction, and willingness to donate again on a five-point ordinal scale. Univariable ordinal logistic regression was used for variable screening, followed by multivariable proportional-odds ordinal logistic regression with purposeful variable selection. Statistical significance was set at *p* < 0.05.

**Results:** The mean donor age was 35.8 ± 11.8 years, and 88% of participants were male. High procedural pain (VAS ≥4) was reported by 73% of donors, whereas 48% expressed high willingness to donate again. After adjustment, procedural pain was the only independent predictor of future donation intention. Each one-point increase in pain score reduced the odds of greater willingness to donate again by 54% (adjusted OR 0.46, 95% CI 0.37–0.57; *p* < 0.001). Donor-perceived needle quality was not associated with willingness to donate again (adjusted OR 1.09, 95% CI 0.47–2.56; *p* = 0.840) and showed no association with procedural pain.

**Conclusions:** Procedural pain, rather than donor-perceived needle quality, was the principal determinant of willingness to donate again. Interventions that improve donor comfort and minimise venepuncture pain may strengthen donor retention. Larger prospective studies using actual donor return behaviour are warranted.

## Introduction

A sufficient and sustainable blood supply depends not only on the recruitment of new donors but also on the retention of existing ones. Repeat voluntary blood donors are considered the foundation of safe blood services because they provide a more predictable blood supply and generally have a lower prevalence of transfusion-transmissible infections than first-time or replacement donors. Consequently, improving donor retention has become a major objective of blood services worldwide, particularly in low- and middle-income countries where blood availability frequently falls short of clinical demand and resources for donor recruitment are limited. (1–3)

The decision to donate blood again is influenced by a complex interaction of individual motivation, previous donation experience, and the quality of care received during donation. While altruism remains an important motivation for first-time donation, the experience during donation largely determines whether an individual returns. Previous studies have shown that donation-related pain, anxiety, vasovagal reactions, bruising, prolonged procedures, and poor communication with collection staff are associated with lower donor satisfaction and reduced donor return. Conversely, donors who experience minimal discomfort and receive supportive care are more likely to become regular voluntary donors. (4–9)

Among the various determinants of donor experience, procedural pain has consistently emerged as one of the strongest predictors of donor satisfaction and future donation behaviour. Pain during venepuncture is associated with greater anxiety, an increased likelihood of vasovagal reactions, and lower rates of repeat donation. (6,8–11) Importantly, procedural pain represents a potentially modifiable aspect of the donation process. Interventions such as improved venepuncture technique, effective donor communication, reassurance before needle insertion, and behavioural strategies to reduce anxiety have all been shown to improve the overall donation experience and may contribute to better donor retention. (9,12)

Despite increasing emphasis on donor-centred care, comparatively little attention has been paid to the characteristics of the blood collection device itself. Blood collection needles are manufactured with differences in bevel geometry, tip sharpness, surface finishing, silicone lubrication, and production quality. Experimental studies in biomedical engineering have demonstrated that these design characteristics influence needle penetration force, tissue deformation, friction during insertion, and the force required to traverse biological tissues. (13–16) Although these mechanical differences may be subtle in clinical practice, they could influence the donor’s perception of insertion comfort and, consequently, the overall donation experience. However, evidence linking donor-perceived needle quality with donor satisfaction or future donation behaviour remains scarce.

This question has practical relevance in Bangladesh, where blood collection systems are procured from multiple manufacturers through different procurement mechanisms. Selection of blood collection bags is generally based on regulatory approval, availability, and cost, whereas donor comfort is rarely considered as a quality indicator during procurement. If donor-perceived needle quality contributes meaningfully to procedural pain or dissatisfaction, improving device quality could represent a simple and cost-effective strategy to enhance donor retention.

Alternatively, if donor willingness is determined primarily by procedural technique and psychological factors rather than needle characteristics, quality improvement efforts should instead focus on staff training, donor communication, and pain reduction during venepuncture.

To our knowledge, no published study has evaluated the relationship between donor-perceived needle quality and future donation intention in Bangladesh. We therefore conducted a cross-sectional analytical study to examine whether donor-perceived needle quality was associated with willingness to donate again among whole-blood donors. We also assessed the contributions of procedural pain, pre-donation fear, vasovagal symptoms, and selected donor- and procedure-related characteristics to future donation intention. We hypothesized that donors reporting poorer perceived needle quality would be less willing to donate again; however, we also sought to determine whether any observed association remained after adjustment for procedural pain and other potential confounding factors.

## Materials and Methods

### Study design and setting

This multicentre cross-sectional analytical study was conducted between **June 2025** and **January 2026** at four tertiary-care hospital blood transfusion services in Bangladesh: Cumilla Medical College Hospital(30), Popular Medical College Hospital(20), Faridpur Medical College Hospital(30), and Dhaka Medical College Hospital(20). The study was designed and reported in accordance with the Strengthening the Reporting of Observational Studies in Epidemiology (STROBE) recommendations for cross-sectional studies.

### Study participants

Eligible participants were consecutive whole-blood donors aged 18–60 years who fulfilled the national eligibility criteria for blood donation and successfully completed whole-blood donation at one of the participating centres during the study period. Donors who declined participation, were unable to provide informed consent, or returned incomplete questionnaires were excluded.

### Sample size and sampling

A total of 100 eligible donors were enrolled using consecutive sampling. Consecutive recruitment was chosen to minimise selection bias and to reflect the routine donor population attending the participating blood centres.

### Data collection

Data were collected immediately after completion of blood donation using a structured interviewer-administered questionnaire developed for this study. The questionnaire consisted of six sections covering demographic characteristics, donation history, pre-donation fear and anxiety, procedural experience, adverse donation symptoms, and future donation intention.

Procedural pain was assessed using a 10-point Visual Analogue Scale (VAS), where 0 represented no pain and 10 represented the worst imaginable pain. Consistent with the predefined study protocol, a VAS score ≥4 was classified as high procedural pain.

Donors were asked to rate their perception of needle quality by indicating whether the needle felt smooth, not smooth, or whether they were unsure. They were also asked whether they perceived the needle as rough during insertion. Additional donor-reported variables included fear before donation, fear after seeing the needle, fear of pain, previous unpleasant donation experience, occurrence of vasovagal symptoms (dizziness, sweating, nausea, weakness, presyncope, or syncope), and overall satisfaction with the donation experience.

Immediately after each donation, the attending phlebotomist independently recorded procedural characteristics, including insertion smoothness, perceived needle sharpness, insertion force, venepuncture attempts, adequacy of blood flow, and professional experience. Donor questionnaires and phlebotomist assessments were completed independently to minimise observer bias.

### Study outcome

The primary outcome was willingness to donate blood again, assessed using a five-point Likert scale ranging from 1 ("very unlikely") to 5 ("very likely"). Because the response categories were inherently ordered, willingness to donate again was analysed as an ordinal outcome.

### Study variables

The primary exposure variable was donor-perceived needle quality. Pre-specified covariates included age, sex, educational attainment, donor type (voluntary, replacement, or family donor), donation status (first-time or repeat donor), procedural pain score, pre-donation fear, previous adverse donation experience, vasovagal symptoms, number of venepuncture attempts, phlebotomist-rated insertion smoothness, needle sharpness, insertion force, and overall donor satisfaction.

### Statistical analysis

Data were entered into Microsoft Excel (Microsoft Corp., Redmond, WA, USA) and analysed using IBM SPSS Statistics version 26.0 (IBM Corp., Armonk, NY, USA).

Continuous variables were summarised as mean ± standard deviation (SD) for normally distributed data or median with interquartile range (IQR) when appropriate. Categorical variables were presented as frequencies and percentages.

Normality of continuous variables was assessed using the Shapiro–Wilk test. Because continuous variables were not normally distributed, non-parametric methods were used for exploratory analyses where appropriate. The primary outcome, willingness to donate again, was analysed using ordinal logistic regression because it was measured on an ordered five-point Likert scale. Mann–Whitney U tests, Kruskal–Wallis tests, and Spearman’s rank correlation were used to explore unadjusted associations where appropriate before multivariable modelling.

The primary analysis used ordinal logistic regression because the outcome variable consisted of ordered categories. Univariable ordinal logistic regression was first performed to identify candidate predictors. Variables with *p* <0.20 were considered for multivariable modelling using a purposeful variable selection strategy. Based on biological plausibility and previous evidence, age, sex, educational level, donor-perceived needle quality, and procedural pain were retained in the final model irrespective of statistical significance. Adjusted odds ratios (aORs) with 95% confidence intervals (CIs) were reported.

The proportional-odds assumption was evaluated using a likelihood-ratio test comparing the proportional-odds model with an unconstrained multinomial logistic regression model. All statistical tests were two-sided, and a *p* value <0.05 was considered statistically significant.

### Ethical considerations

The study protocol was reviewed and approved by the Institutional Review Board of Cumilla Medical College Hospital (Approval No. **June25-03)** Administrative permission was obtained from each participating institution before data collection. Written informed consent was obtained from all participants prior to enrolment. Participation was voluntary, and donors were informed that refusal or withdrawal would not affect their eligibility for blood donation or future medical care. All data were anonymised before analysis and handled confidentially in accordance with the principles of the Declaration of Helsinki.

## Results

### Participant characteristics

A total of 100 whole-blood donors were included. The mean age was 35.8 ± 11.8 years (median 34.5, IQR 25.0–46.5), and the sample was predominantly male (88, 88.0%; female 12, 12.0%). Educational attainment ranged from primary (22, 22.0%) to postgraduate (15, 15.0%), with secondary, HSC, and graduate levels accounting for 18 (18.0%), 25 (25.0%), and 20 (20.0%), respectively. Occupations were diverse, the most common being engineer (18, 18.0%), service (15, 15.0%), and teacher (15, 15.0%). Half of the donors were voluntary (50, 50.0%), 40 (40.0%) were replacement donors, and 10 (10.0%) were family donors. Repeat donors slightly outnumbered first-time donors (57, 57.0% vs 43, 43.0%), with a mean of 3.2 ± 3.7 previous donations (median 2.0, IQR 0.0–6.0).

### Donation experience

Perceived procedural pain was substantial: the mean Pain VAS score was 5.4, and 73 (73.0%) donors reported high pain (VAS ≥4). Pre-donation fear was broadly distributed across the 1–5 scale for fear before donation, fear on seeing the needle, and fear of pain, with no single category predominating. The needle was perceived as smooth by 60 (60.0%) donors, not smooth by 28 (28.0%), and 12 (12.0%) were unsure; 28 (28.0%) explicitly described the needle as rough. Multiple venepuncture attempts occurred in 9 (9.0%) donors, blood flow was rated good in 72 (72.0%), and haematoma and premature cessation of donation were uncommon (5, 5.0% and 3, 3.0%, respectively).

At least one vasovagal symptom was reported by 54 (54.0%) donors. The most frequent individual symptoms were weakness (24, 24.0%) and dizziness (17, 17.0%), followed by sweating (12, 12.0%) and nausea (8, 8.0%); fainting sensation was rare (1, 1.0%) and no donor experienced syncope (0, 0.0%). On phlebotomist-rated measures, insertion was characterised as smooth in 64 (64.0%), sharpness as good in 62 (62.0%), and insertion force as low in 58 (58.0%); phlebotomist experience was evenly distributed across categories. Overall satisfaction was high (ratings 4–5) in 50 (50.0%) donors.

### Willingness to donate again

Willingness to donate again spanned the full 1–5 scale: 12 (12.0%) responded 1, 13 (13.0%) responded 2, 27 (27.0%) responded 3, 20 (20.0%) responded 4, and 28 (28.0%) responded 5. Overall, 48 (48.0%) expressed higher willingness (4–5), 27 (27.0%) were neutral, and 25 (25.0%) expressed low willingness (1–2) (Figure 2).

### Univariable screening

All continuous variables departed significantly from normality on Shapiro–Wilk testing (all p < 0.001). In the univariable ordinal logistic screen, perceived pain (Pain VAS) was strongly associated with willingness to donate again (p < 0.001), and insertion force met the pre-specified screening threshold (p = 0.140). All other candidate variables fell below the p < 0.20 threshold, including needle roughness (p = 0.927), the pre-donation fear items (p = 0.273–0.914), previous bad experience (p = 0.333), vasovagal reaction (p = 0.816), donation status (p = 0.406), multiple attempts (p = 0.650), and phlebotomist-rated smoothness (p = 0.816), sharpness (p = 0.742), and experience (p = 0.746). Pain VAS and insertion force advanced to the multivariable model; age (p = 0.189), sex (p = 0.243), and education (p = 0.311) were retained a priori as covariates (Table 2).

**Table 1.** Characteristics of blood donors (N = 100)

| **Characteristic** | **N = 100** |
| --- | --- |
| **Age, years — mean ± SD (median [IQR])** | 35.8 ± 11.8 (34.5 [25.0–46.5]) |
| **Sex** |  |
| Male | 88 (88.0) |
| Female | 12 (12.0) |
| **Education** |  |
| Primary | 22 (22.0) |
| Secondary | 18 (18.0) |
| HSC | 25 (25.0) |
| Graduate | 20 (20.0) |
| Postgraduate | 15 (15.0) |
| **Donor type** |  |
| Voluntary | 50 (50.0) |
| Replacement | 40 (40.0) |
| Family | 10 (10.0) |
| **Donation status** |  |
| First-time | 43 (43.0) |
| Repeat | 57 (57.0) |
| **Previous donations — mean ± SD (median [IQR])** | 3.2 ± 3.7 (2.0 [0.0–6.0]) |
| **Pain VAS (0–10) — mean** | 5.4 |
| **High pain (VAS ≥4)** | 73 (73.0) |
| **Needle perceived smooth** |  |
| Yes | 60 (60.0) |
| No | 28 (28.0) |
| Unsure | 12 (12.0) |
| **Needle perceived rough — Yes** | 28 (28.0) |
| **Multiple venepuncture attempt — Yes** | 9 (9.0) |
| **Blood flow good** | 72 (72.0) |
| **Haematoma — Yes** | 5 (5.0) |
| **Donation stopped — Yes** | 3 (3.0) |
| **Any vasovagal symptom (composite)** | 54 (54.0) |
| Weakness | 24 (24.0) |
| Dizziness | 17 (17.0) |
| Sweating | 12 (12.0) |
| Nausea | 8 (8.0) |
| Fainting sensation | 1 (1.0) |
| Syncope | 0 (0.0) |
| **Phlebotomist-rated insertion smooth** | 64 (64.0) |
| **Needle sharpness good** | 62 (62.0) |
| **Insertion force low** | 58 (58.0) |
| **Overall satisfaction high (4–5)** | 50 (50.0) |

Values are n (%) unless otherwise indicated. HSC, Higher Secondary Certificate; VAS, visual analogue scale. Vasovagal composite = any of dizziness, sweating, nausea, weakness, fainting sensation, or syncope.

**Table 2.**
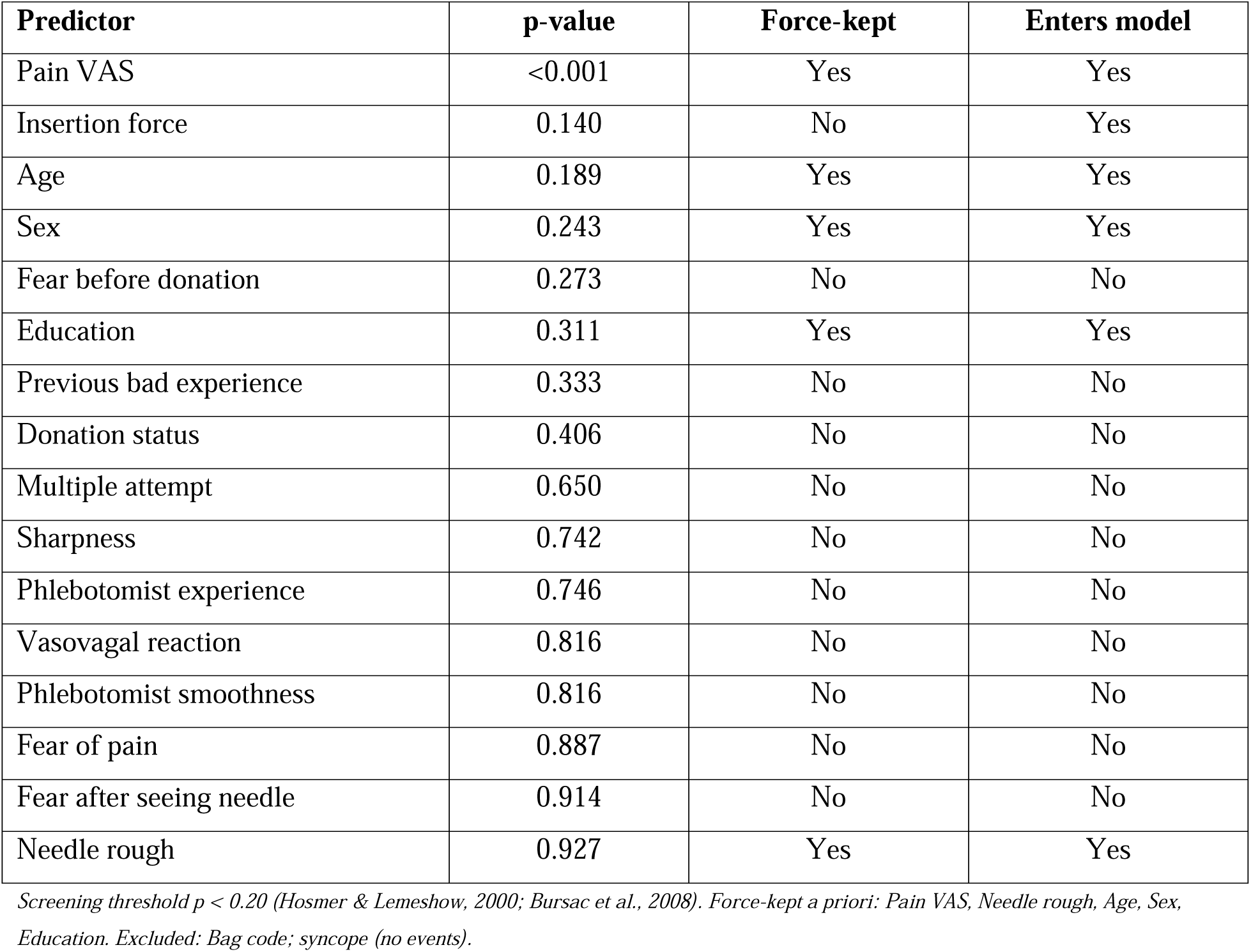
Univariable screen (ordinal logistic regression, likelihood-ratio test)

### Perceived needle quality and pain

Because perceived needle quality was the primary exposure of interest, its relationship with both the outcome and pain was examined directly. Donor-perceived needle smoothness (smooth/not smooth/unsure) was not associated with willingness to donate again (likelihood-ratio χ² = 0.04, df = 2, p = 0.98; Kruskal–Wallis p = 0.98), with near-identical mean willingness across groups (3.4, 3.4, and 3.5, respectively). Perceived needle quality was likewise unrelated to procedural pain: Pain VAS did not differ between donors who described the needle as rough versus not rough (median 6 vs 5; Mann–Whitney p = 0.84) or across smoothness categories (Kruskal– Wallis p = 0.91; Spearman ρ = −0.03). Thus, although pain was strongly associated with retention intention, it did not appear to be explained by donor-perceived needle quality.

### Multivariable ordinal logistic regression

The proportional-odds assumption was supported by a likelihood-ratio test comparing the proportional-odds model with an unconstrained multinomial model (χ² = 15.79, df = 21, p = 0.78). In the adjusted model, higher perceived pain was independently associated with lower willingness to donate again: each one-point increase on the VAS reduced the odds of being in a higher willingness category by approximately 54% (adjusted OR 0.46, 95% CI 0.37–0.57; p < 0.001). No other variable was independently associated with the outcome — insertion force (moderate vs low OR 1.00, 95% CI 0.42–2.39; high vs low OR 2.29, 95% CI 0.63–8.30), needle roughness (OR 1.09, 95% CI 0.47–2.56), age (OR 1.00, 95% CI 0.97–1.03), female sex (OR 1.78, 95% CI 0.54–5.92), and education above secondary level (OR 1.40, 95% CI 0.64–3.06) all had confidence intervals that included 1 (Table 3, Figure 1).

**Figure 1.**
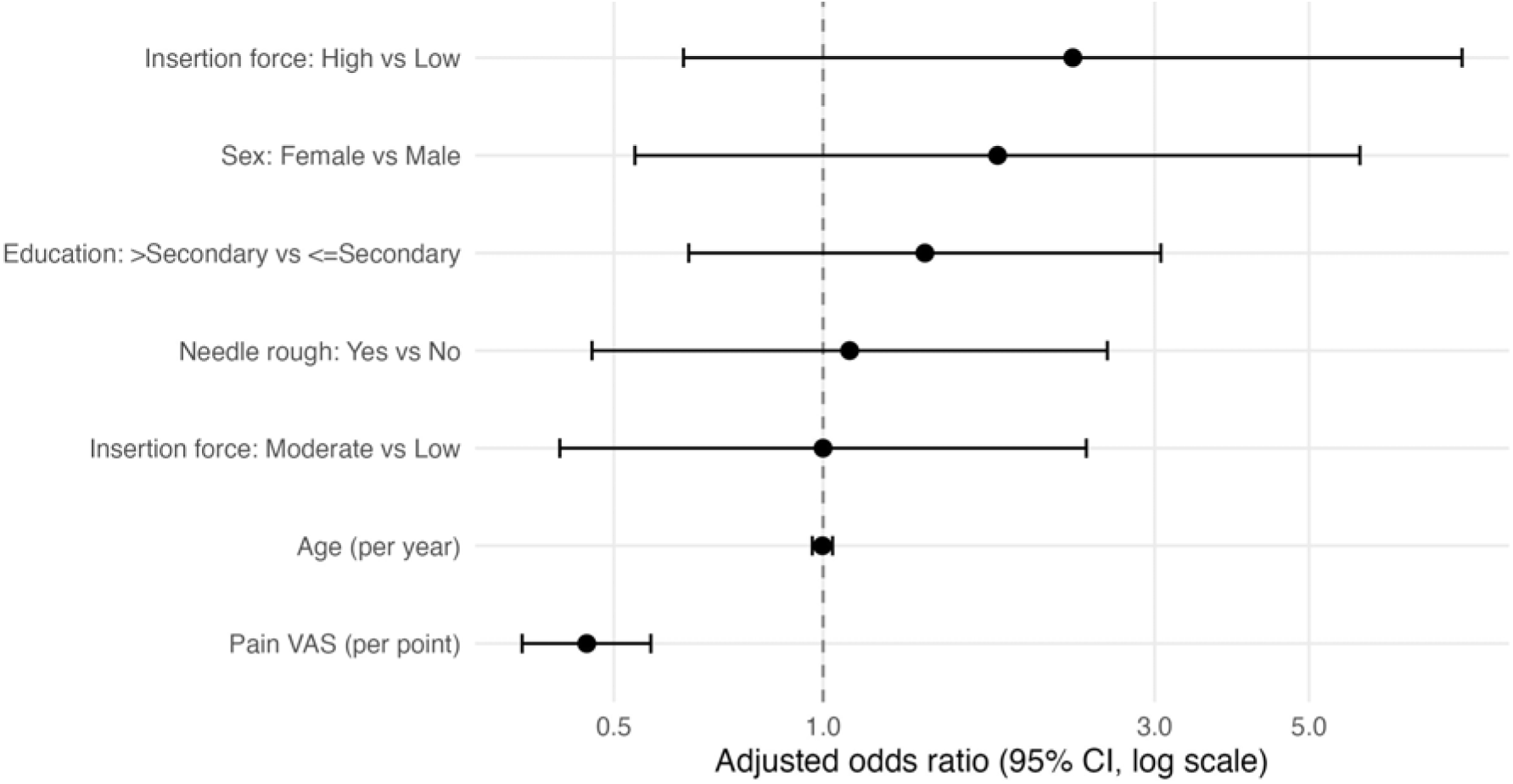
Adjusted odds ratios (95% CI) for willingness to donate again, from the multivariable ordinal logistic regression model.

**Figure 2.**
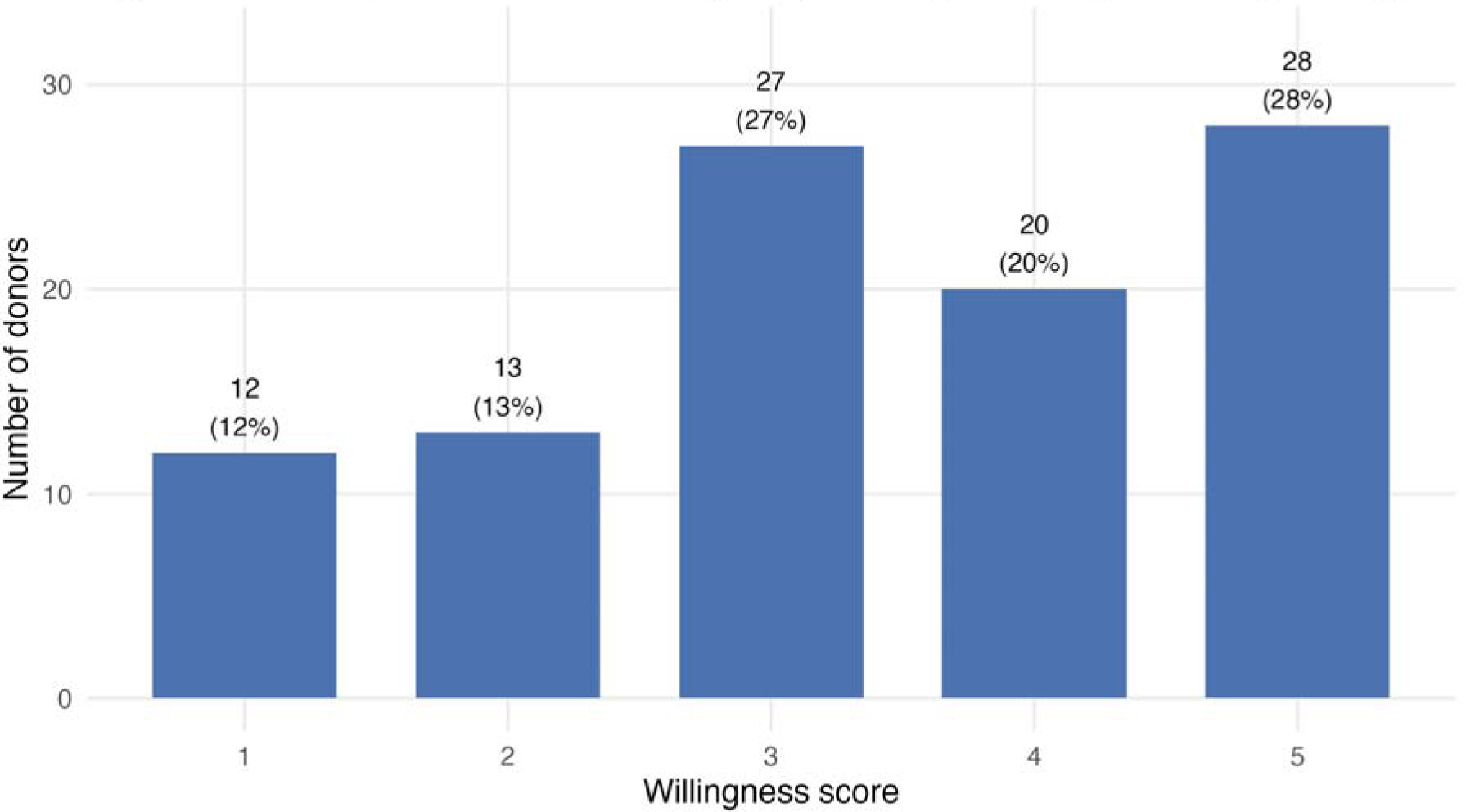
Distribution of willingness to donate again (1 = very unlikely to 5 = very likely).

**Table 3.** Multivariable ordinal logistic regression.

| Predictor | Adjusted OR | 95% CI | p-value |
| --- | --- | --- | --- |
| Pain VAS (per point) | 0.46 | 0.37–0.57 | <0.001 |
| Insertion force: Moderate vs Low | 1.00 | 0.42–2.39 | 1.000 |
| Insertion force: High vs Low | 2.29 | 0.63–8.30 | 0.209 |
| Needle rough: Yes vs No | 1.09 | 0.47–2.56 | 0.840 |
| Age (per year) | 1.00 | 0.97–1.03 | 0.910 |
| Sex: Female vs Male | 1.78 | 0.54–5.92 | 0.346 |
| Education: >Secondary vs | 1.40 | 0.64–3.06 | 0.398 |
| ≤Secondary |  |  |  |

Outcome: willingness to donate again (1–5, ordered). OR > 1 indicates higher odds of greater willingness. Two-stage purposeful selection (screen $p < 0.20$ ; Pain VAS, Needle rough, Age, Sex, Education force-kept). Proportional-odds assumption checked by likelihood-ratio test (polr vs multinomial). CI, confidence interval.

## Discussion

In this cross-sectional study of 100 whole-blood donors, perceived procedural pain emerged as the single dominant correlate of the intention to donate again. After adjustment, each one-point increase on the 0–10 visual analogue scale was associated with an approximately 54% reduction in the odds of expressing greater willingness to return (adjusted OR 0.46, 95% CI 0.37–0.57). By contrast, the study’s named exposure of interest — donor-perceived needle quality — was not associated with retention intention, and showed no relationship with the pain that donors reported. Pre-donation fear, vasovagal symptoms, donation status, and phlebotomist-related ratings were likewise not independently associated with the outcome. Taken together, these findings reposition procedural pain, rather than the physical characteristics of the needle, as the most actionable target for protecting donor return in this setting.

### Pain as a modifiable determinant of donor return

The strength and consistency of the pain–retention association is the central finding of this study. The graded relationship across the willingness categories (Figure 3), with median pain falling steadily from the least to the most willing donors, is consistent with a growing body of work identifying venepuncture pain as a determinant of the donation experience. In a two-centre, individually randomised controlled trial of Australian donors, Gilchrist and colleagues found that venepuncture pain was associated with vasovagal reactions, lower satisfaction, and a reduced likelihood of returning within six months, and that donor fear predicted higher pain even after accounting for estimated blood volume, age, sex, and donation experience [17]. More recently, analyses from the large STRIDES cohort in England reported that venepuncture pain and donation anxiety mediated the relationship between donation history and vasovagal symptom reporting, reinforcing the role of pain as a mechanism rather than an incidental by-product of donation [18]. The present results extend this literature to a South Asian, predominantly voluntary- and replacement-donor population, and suggest that pain is associated not only with adverse physiological reactions but directly with a donor’s stated intention to return.

**Figure 3.**
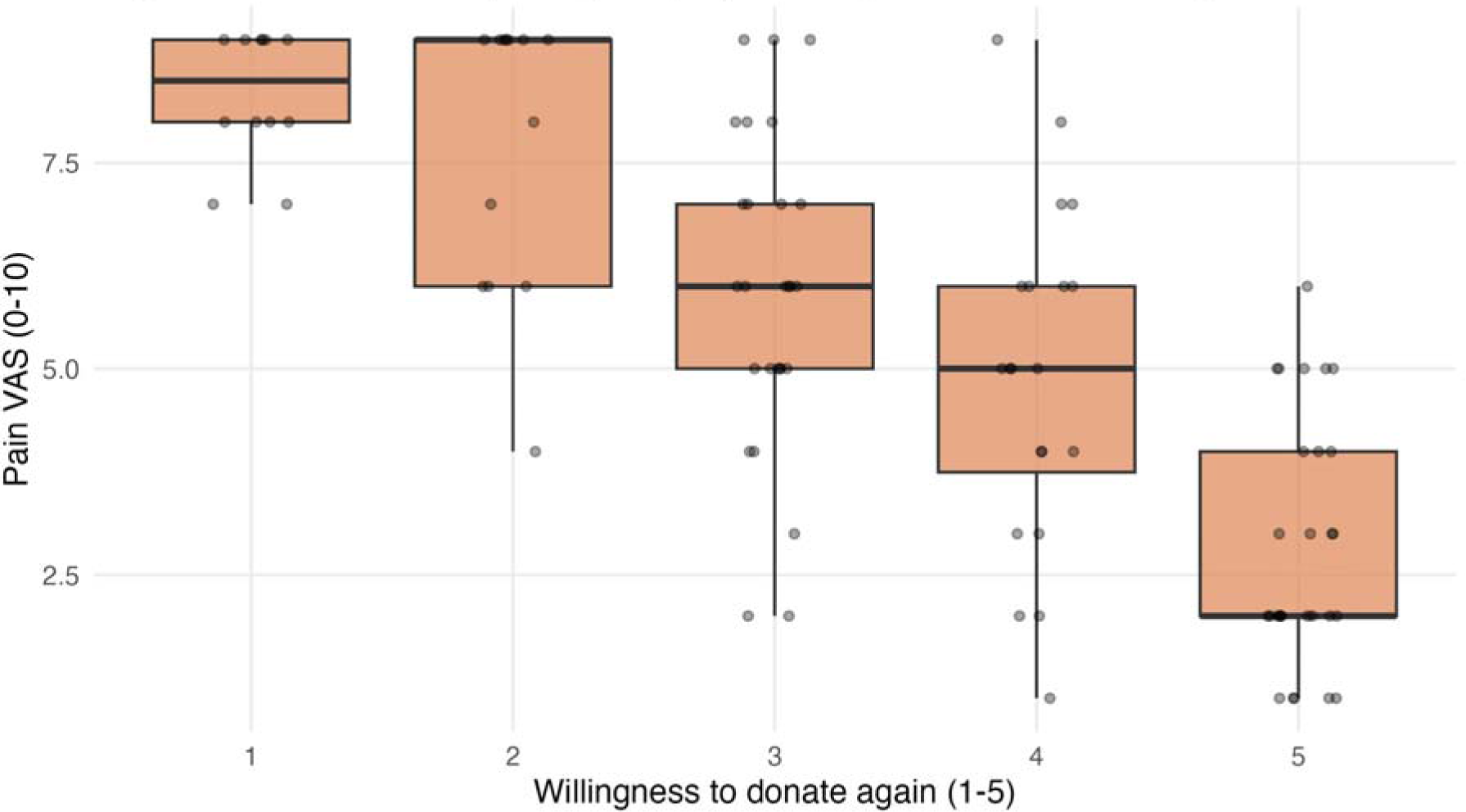
Procedural pain (VAS) by willingness-to-donate-again category.

Crucially, pain is modifiable. Interventions addressing the painfulness of the donation experience — including distraction and coping techniques, applied muscle tension, and supportive communication from collection staff — have been shown to improve aspects of the donor experience. Stewart and colleagues demonstrated that phlebotomist interpersonal skill predicts a reduction in donor reactions, underscoring that the human and procedural context of venepuncture, not merely the device, shapes how donation is experienced [19]. Framed this way, the findings argue for embedding pain assessment and pain-reduction strategies into routine donor care as a retention measure, particularly for donors who report high pain at their index donation.

### Perceived needle quality and the donation experience

Contrary to the study hypothesis, donor-perceived needle quality was not associated with willingness to donate again. Neither the donor’s judgement of needle smoothness nor an explicit description of the needle as rough predicted the outcome, and these perceptions were unrelated to the level of pain reported. This dissociation is informative: it suggests that, at least as perceived and reported by donors, the physical needle is not the principal driver of the painful or aversive elements of donation in this population. Pain appears to arise from factors that donors do not necessarily attribute to the needle itself — which may include venepuncture technique, vein selection, anxiety-amplified pain perception, and individual pain sensitivity.

This result should be interpreted with appropriate caution rather than as definitive evidence that needle characteristics are irrelevant. Donor-reported smoothness is a subjective, single-item perception measured after the fact, and may correlate only weakly with objective needle properties such as gauge, bevel geometry, or coating. It is therefore possible that genuine differences in needle quality exist but are not reliably perceived or recalled by donors. Nonetheless, from a programmatic standpoint, the absence of any association between perceived needle quality and either pain or retention tempers the expectation that procurement decisions centred on needle product alone will materially improve donor return, and redirects attention toward the procedural and psychological management of pain.

### Fear, vasovagal symptoms, and donor characteristics

Pre-donation fear items were not independently associated with willingness to donate again in this sample. This contrasts with longitudinal work by France and colleagues, who reported that donation-related fears predict vasovagal reactions and attrition among high-school donors [20], and that fear is associated with attrition of first-time donors through lower donation confidence and more negative attitudes [21]. It also contrasts with cross-sectional findings from Saqlain and colleagues in Lahore, Pakistan, who observed that blood-donation fears were inversely related to willingness for future donation and diminished with greater donation experience, higher education, and older age [23]. A plausible explanation for the weaker fear signal here is the composition of the sample: more than half of participants were repeat donors, a group in whom fear tends to be attenuated through habituation. In more experienced donors, the proximal sensory experience of pain may carry greater weight for future intention than anticipatory fear.

Vasovagal symptoms were common, reported by just over half of donors, but were not independently associated with retention intention, and frank syncope did not occur. Although vasovagal reactions are a well-established risk factor for reduced donor return — as shown by Olatunji and colleagues [22] and in the first-time-versus-repeat cohort analysis of Wiersum-Osselton and colleagues [24] — most symptoms in this sample were mild (predominantly weakness and dizziness), which may explain their limited influence on stated intention. Donation status, multiple venepuncture attempts, and phlebotomist-rated measures were similarly non-significant; the wide confidence intervals around several categorical estimates indicate that the study was not powered to detect modest effects for these less prevalent exposures.

### Implications for donor management

The practical message of this study is straightforward: managing pain is likely to be more productive for donor retention than focusing on needle-product specifications. Blood centres in comparable settings could incorporate brief pain assessment into the post-donation interaction, identify donors reporting high pain for targeted follow-up and reassurance, train collection staff in both technical venepuncture skill and supportive communication, and evaluate low-cost pain-mitigation strategies. Because the donation experience is increasingly understood as a service encounter shaped by both cognitive appraisal and emotion — as Martín-Santana and colleagues showed in their analysis of service quality, anticipated emotions, and donor loyalty in Spanish blood centres [25] — pain reduction may exert effects on return that extend beyond the physical sensation to the donor’s overall evaluation of the experience.

### Strengths and limitations

Strengths of this study include the use of a pre-specified two-stage modelling strategy with purposeful variable selection [26,27], an ordinal regression approach that preserved the full response scale [28], and verification of the proportional-odds assumption. The direct interrogation of the study’s primary exposure against both the outcome and the proposed mediating pathway adds analytic transparency.

Several limitations should be acknowledged. First, the sample of 100 donors limited statistical power, producing wide confidence intervals for less prevalent categorical predictors and precluding detection of modest associations; the model was adequately specified (approximately 14 observations per estimated coefficient), but small-sample imprecision remains. Second, the outcome was a single-item, five-point measure of donation intention rather than verified return behaviour, and intention is an imperfect proxy for subsequent action. Third, the sample was overwhelmingly male (88%), which constrains generalisability — particularly given evidence that female donors experience higher rates of needle-related complications [24] — and may have limited the ability to detect sex-related effects. Fourth, perceived needle quality and pain were self-reported and subject to recall and reporting biases. Finally, the cross-sectional, single-time-point design precludes causal inference; longitudinal studies linking the index donation experience to actual return are needed to confirm whether reducing procedural pain translates into measurable gains in donor retention.

## Conclusion

In this multicentre cross-sectional study, procedural pain emerged as the strongest independent factor associated with donors’ willingness to donate again, whereas donor-perceived needle quality was not independently associated with future donation intention or procedural pain. These findings suggest that the overall donation experience is influenced more by how the donation is performed than by donors’ perceptions of the collection needle itself.

Improving venepuncture technique, strengthening staff communication, and adopting donor-centred approaches to minimise procedural discomfort may represent practical and achievable strategies for enhancing the donation experience and encouraging repeat blood donation. Although the physical characteristics of blood collection devices remain important for quality assurance and procedural safety, our findings indicate that optimising donor care may have a greater impact on future donation intention than focusing solely on perceived needle quality.

Because this study assessed intended rather than actual donation behaviour and included a relatively modest sample size, larger multicentre prospective studies with longitudinal follow-up are needed to determine whether interventions that reduce procedural pain translate into improved donor retention. Such evidence would help guide donor-centred quality improvement initiatives and support policies aimed at strengthening voluntary blood donation programmes.

## Data Availability

All data produced in the present study are available upon reasonable request to the authors

## Notes

### Competing Interest Statement

The authors have declared no competing interest.

### Author Declarations

Ethical review board of Comilla Medical College. Ref no- CoMC/ERB/02

